# High-dimensional profiling of pediatric immune responses to solid organ transplantation

**DOI:** 10.1101/2022.08.17.22278895

**Authors:** Mahil Rao, Meelad Amouzgar, James T. Harden, M. Gay Lapasaran, Amber Trickey, Brian Armstrong, Jonah Odim, Tracia Debnam, Carlos O. Esquivel, Sean C. Bendall, Olivia M. Martinez, Sheri M. Krams

## Abstract

Solid organ transplant remains a life-saving therapy for children with end-stage heart, lung, liver, or kidney disease; however, ∼25% of allograft recipients experience acute rejection within the first 12 months after transplant. Our ability to detect rejection early and to develop less toxic immunosuppressive agents is hampered by an incomplete understanding of the immune changes associated with rejection, particularly in the pediatric population. Here we used high-dimensional single-cell proteomic technologies (CyTOF) to generate the first detailed, multi-lineage analysis of the peripheral blood immune composition of pediatric solid organ transplant recipients. We report that the organ transplanted impacts the immune composition post-transplant. When taking these allograft-specific differences into account, we further observed that differences in the proportion of subsets of CD8 and CD4 T cells were significantly associated with allograft health. Together, these data form the basis for mechanistic studies into the pathobiology of rejection to develop less invasive tools to identify early rejection and new immunosuppressive agents with greater specificity and less toxicity.

## Introduction

For children with end-stage heart, liver, kidney, or lung disease, solid organ transplant confers a significant benefit to both quality and quantity of life. Nearly 3000 children underwent single organ transplant in 2021 in the United States.^1^ Receiving an allograft confers a new set of challenges, however, as the recipient will also need lifelong immunosuppression to prevent the recipient’s immune system from attacking the donor organ. Maintaining an appropriate degree of immunosuppression remains one of the major challenges in transplant medicine today: too little immunosuppression and the recipient’s immune system will attack the donor organ, leading to acute rejection, while too much immunosuppression leaves the recipient unnecessarily vulnerable to opportunistic infections,^2^ organ damage,^3^ and post-transplant lymphoproliferative disorder.^4^ It is estimated that between 25% and 40% of solid organ transplant recipients will have at least one episode of acute rejection in the first 12 months after transplant.^5–8^

The alloimmune response involves both innate and adaptive components and the cellular populations involved in rejection has been the subject of several excellent recent reviews.^9–13^ Much of the available data is based on analysis of single lineages, single allograft type, or is taken from adult patients. Here we addressed this gap in the literature by performing a detailed, multi-lineage, single-cell analysis of the peripheral blood immune composition of pediatric solid organ transplant recipients with modern high-dimensional proteomic technologies (CyTOF). We constructed immune profiles from pediatric heart, liver, kidney, and small intestine recipients at various times after transplant with either stable graft function or experiencing rejection by measuring the proportion of 29 subpopulations of peripheral blood mononuclear cells. Using a combination of supervised and unsupervised analyses, we define the immune features associated with allograft type and the distinct changes in immune composition associated with allograft rejection.

## Methods

### Patient samples

Patient samples were obtained from a biobank generated as a part of the Clinical Trials in Organ Transplantation in Children-06 (CTOTC-06) study.^14^ CTOTC-06 was an NIH-sponsored prospective, observational study in which 944 pediatric subjects (neonate to 21 years of age) were enrolled pre-transplant and post-transplant at seven clinical sites. Institutional Review Board (IRB) approval was obtained, and all procedures were performed in accordance with the relevant guidelines and regulations. For participants under the age of 18 years, informed consent was obtained from a parent and/or legal guardian. Of these subjects, 872 received transplants. Immunosuppressive and anti-viral therapy were per each center’s standard protocol. Prospective blood samples were collected (n=4753) at enrollment or transplant, every 3 months during the first 2 years and twice yearly thereafter, regardless of allograft health. Follow up was for a minimum of 12 months and a maximum of four years based on the time of enrollment. Subjects enrolled post-transplant were within three years of transplantation. Peripheral blood mononuclear cells (PBMCs) were isolated from these samples and cryopreserved.

In this study, a total of 52 samples were profiled **(Table 1)**. We based our power calculations on data from a 2016 study using a similar approach in a different cohort of pediatric liver transplant recipients.^15^ In that study, we observed more than a two-fold difference in the percentage of populations of interest between patient groups. Assuming a normal distribution and type I error rate=0.05, two-sided Satterthwaite’s t-tests for unequal variance were estimated to have >99% power to detect similar differences between rejection and stable patients within each organ group. Twenty-four of the samples - the “stable” cohort - were from patients who had no biochemical or biopsy evidence of rejection or infection in the 12 months before and the 12 months after sample collection; the remaining 28 samples – the “rejection” cohort - were from patients who had biopsy-proven rejection in the 30-day period after the sample was collected. Of note, no two samples used in this study were collected from the same patient. Cryopreserved PBMCs were thawed and barcoded with palladium isotopes. Use of palladium-isotope barcoding permits pooling of the sample prior to staining, thereby reducing sample-to-sample data variability from slight differences in staining time, antibody concentration, etc. Samples were then pooled and stained with a panel of 37 metal-conjugated antibodies against cell surface proteins designed to identify specific immune cell lineages. The pooled sample was then subject to analysis on a mass cytometer. The mass cytometer data were normalized and debarcoded. The data from the patient samples were collected over the course of three experiments; to account for subtle differences in staining patterns and variability in machine conditions, single aliquots from the same two healthy donors were included with each run alongside the patient samples; these are hereafter referred to as “anchor” samples. Batch correction was performed with the anchor samples included in the experiments using the approach described in Schuyler, et al.^16^

**Table 1:** Characteristics of patient samples used in the study

|  | Stable | Rejection | p value |
| --- | --- | --- | --- |
| Total Patients | 24 (46%) | 28 (54%) | - |
| Male Sex | 14 (58%) | 16 (57%) | - |
| Heart Recipient | 7 (29%) | 11 (39%) | - |
| Liver Recipient | 12 (50%) | 9 (32%) | - |
| Kidney Recipient | 4 (17%) | 7 (25%) | - |
| Intestine Recipient | 1 (4%) | 1 (4%) | - |
| Age at Transplant (years) | 6.9 ± 6.8 | 8.7 ± 5.8 | 0.302 |
| Age at Collection (years) | 9.3 ± 6.3 | 9.9 ± 6.0 | 0.710 |
| Time Post Transplant at Collection (days) | 878 ± 446 | 448 ± 450 | 0.001 |
| Time Between Collection and Rejection Diagnosis (days) | N/A | 6.1 ± 9.5 | - |

### Reagents

All CyTOF reagents and antibodies (Table 2) were purchased from Standard Biotools (South San Francisco, CA, USA), except as noted. Cell-ID cisplatin (Standard Biotools) was used as per manufacturer’s directions to differentiate live vs dead populations. Monoclonal antibodies against NKG2C and CD5 were purchased from R&D Systems (Minneapolis, MN, USA) and Biolegend (San Diego, CA, USA), respectively, and were conjugated to various heavy metals using Maxpar Antibody Labeling Kits from Standard Biotools.

**Table 2:**
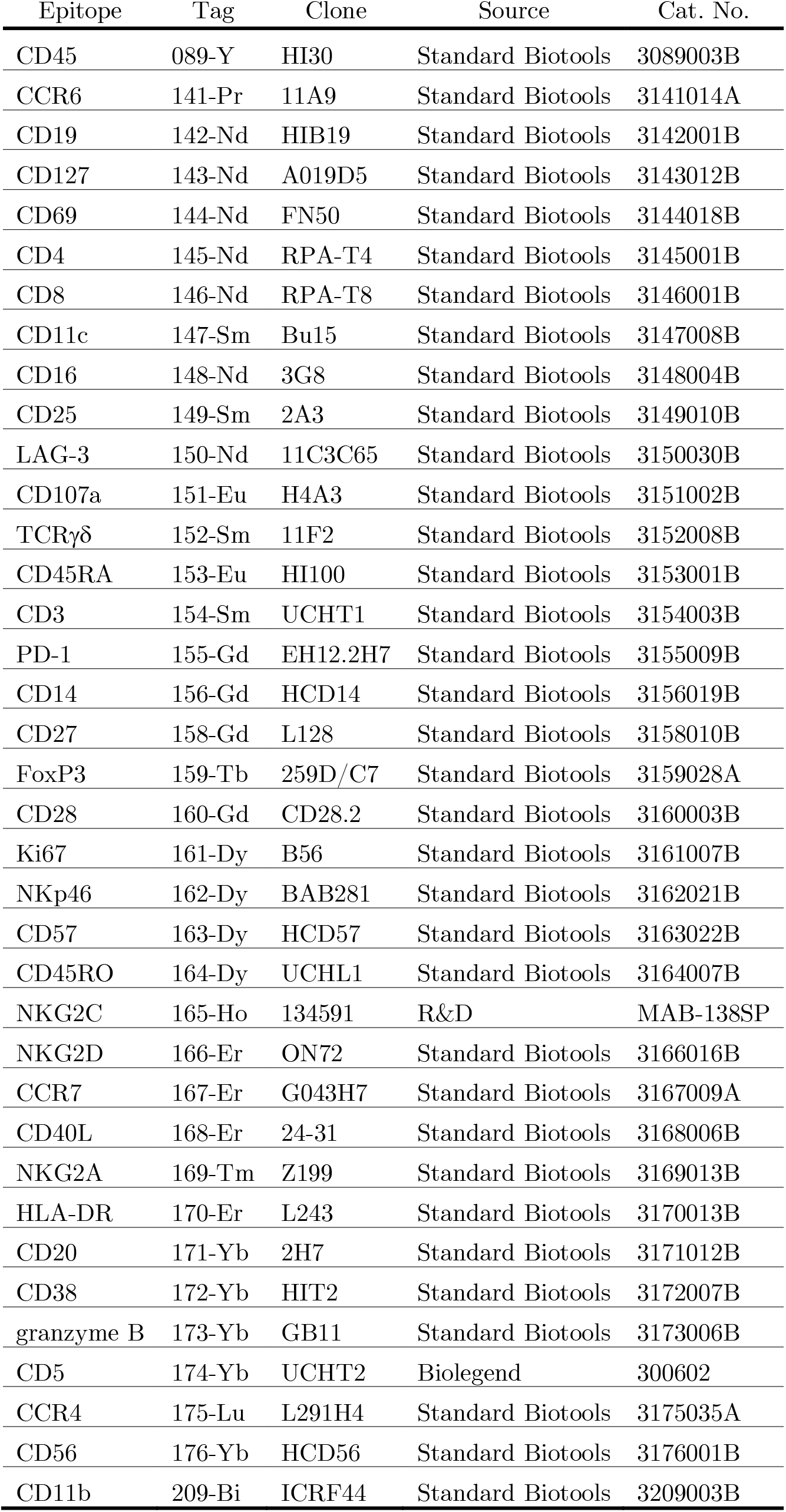
Antibodies used for CyTOF staining

### CyTOF staining

PBMCs from patient samples or anchor controls were thawed into warmed Roswell Park Memorial Institute 1640 media (RPMI) with 20% fetal bovine serum, washed once with phosphate-buffered saline (PBS), and stained with 1 ml of 0.25 mM cisplatin (Standard Biotools) for 5 minutes at room temperature for exclusion of dead cells. Samples were then washed with Cell Staining Buffer (CSB) (Standard Biotools) and barcoded using a Cell-ID™ 20-Plex Pd Barcoding Kit (Standard Biotools) of lanthanide-tagged cell-reactive metal chelators that will covalently label samples with a unique combination of palladium isotopes according to the manufacturer’s instructions. Samples were washed twice with CSB, combined, and incubated with Fc block (Biolegend) for 10 minutes at room temperature. Cells were then stained with a heavy metal-labeled monoclonal antibody cocktail directed against cell surface markers in CSB for 30 minutes at room temperature. Cells were washed twice with FoxP3 permeabilization buffer (Invitrogen) and incubated in FoxP3 permeabilization buffer with an intracellular staining cocktail for 30 minutes at room temperature. Cells were twice washed with CSB, then incubated overnight at 4°C with 1 mL of 125 nM iridium intercalator (Standard Biotools) in Fix/Perm Buffer for DNA staining. Cells were washed twice with CSB and washed twice with cell acquisition solution (CAS, Standard Biotools), then passed through a 50-μm filter and analyzed on a Helios mass cytometer (250–450 events per second).

### CyTOF data pre-processing

Flow cytometry standard (FCS) files were concatenated, normalized (using beads) and debarcoded using the Helios software. Normalized and debarcoded data was gated using CellEngine (http://cellengine.com). Briefly, cellular events were identified using a DNA-intercalator dye (191-iridium and 193-iridium double positive). Singlets were extracted from all cellular events based on event length, and live cells were extracted from singlets based on cisplatin staining (195-cisplatin negative). Exported FCS files of the raw counts of all markers and cell annotations were imported into R using FlowCore.^17^ Cells were annotated with their most confident, terminally-defined manual gate in CellEngine. If cells were unable to be annotated by a terminal gate (e.g., memory B cell), they were annotated by the closest upstream gate (e.g., B cell). We removed duplicate cells from upstream FCS files but retained the most terminal gate for each cell to obtain a single-cell matrix of cells and features that includes all live cells gated within the T cell, B cell, and myeloid populations. Each marker was transformed using an arcsinh scale of 5 and percentile normalized using a quantile value of 0.999 across all cells. Cell proportions were computed based on the major lineage they occupy: T cells, B cells, NK cells or CD56-myeloid cells.

### Technical effect discovery and correction

We implemented linear mixed models to unveil drivers of clinical and biological variation using the *VariancePartition* package.^18^ We identified drivers of variation by quantifying the variation in each trait that is attributable to differences in metadata such as cell type, sample, age at collection, allograft tissue, and allograft health. We input our single-cell matrix of cells and markers, and modeled cell-level metadata. The contribution of each variable is in terms of the fraction of variation explained (FVE). Mixed modeling and principal component analysis (PCA) unveiled that sample-level heterogeneity was responsible for the largest variation in the dataset after cell-type signals. Due to technical effects from separate CyTOF runs on patient samples, we implemented batch correction using the CyTOF BatchAdjust algorithm on raw data to harmonize all patient samples. Anchor samples included in each batch were used to compute adjustment factors for each channel in each batch.^16^ Batch-corrected data was imported into CellEngine, manually gated, and then imported into R for preprocessing and transformation prior to further analysis.

### Statistics

Correlations between marker expression values were generated using the Pearson correlation method. Statistical testing of independent groups was performed using the Wilcoxon Rank Sum Test. Where applicable, multiple hypothesis correction was performed using the false discovery rate. Aggregate marker expression values were computed using the median to maintain robustness to outliers. We used logistic regression with the generalized linear model (glm) function in the *stats* package of R to compare between patient samples.

### Dimensionality Reduction

We used four dimensionality-reduction algorithms for analysis: two unsupervised methods - principal component analysis (PCA) and uniform manifold approximation and projection (UMAP) visualization - and two supervised methods – linear discriminant analysis (LDA) and sparse linear discriminant analysis (sparseLDA). Data was z-scaled and centered prior to PCA transformation, and PCA was done in R using *prcomp* in the *stats* package.^19^ UMAP visualization was performed using the *uwot* package.^19^ UMAP parameter tuning was done using the alpha and beta parameters. For the single-cell UMAP, we visualized only cells in the terminally gated populations. LDA was performed using the *MASS* and hsslda packages in R.^20–22^ Data was z-scaled and centered prior to LDA to improve feature interpretability. Feature importance was inferred using the magnitude and direction of the LD coefficient with respect to the respective class distribution in linear discriminant space. Permutation analysis of the allograft versus randomized groups was done using *sparseLDA* in R.

### Supervised Analysis with Manual Gating

Cells were gated into 29 populations based on surface and intracellular marker staining patterns. The abundance of each population as a percentage of the parent lineage (T cell, B cell, NK cell, CD56-myeloid cell) was computed. Comprehensive comparisons between CyTOF and flow cytometric data have been described by Bendall, et al.^23^ Plots were constructed using either Prism 9.0 (Graphpad Software) or the various R packages listed. Comparisons between groups were performed using the statistical test and the assumptions given in the legend below each figure. The number of asterisks indicates the p-value obtained from the appropriate statistical test (* = p < 0.05; ** = p < 0.001; *** = p < 0.0005.)

## Results

### Study Population and Construction of Immune Profiles

**Figure 1A** shows the experimental approach used for this study. Immune profiles were constructed from PBMCs from 52 pediatric transplant recipients as described in *Methods*. The demographics and clinical characteristics of the patients are shown in **Table 1**. The “stable” and “rejection” cohorts were demographically similar: 46% were male; 29% were heart recipients, 50% were liver recipients, and 17% were kidney recipients. Of note, we required 12 months of stability before sample collection in our “stable” cohort to minimize the effects of treatment for a prior rejection episode; therefore, the time between transplant and sample collection was significantly greater in the “stable” cohort compared to the “rejection” cohort. Clinical samples were processed as described in the methods and **Figure 1B**.

**Figure 1:**
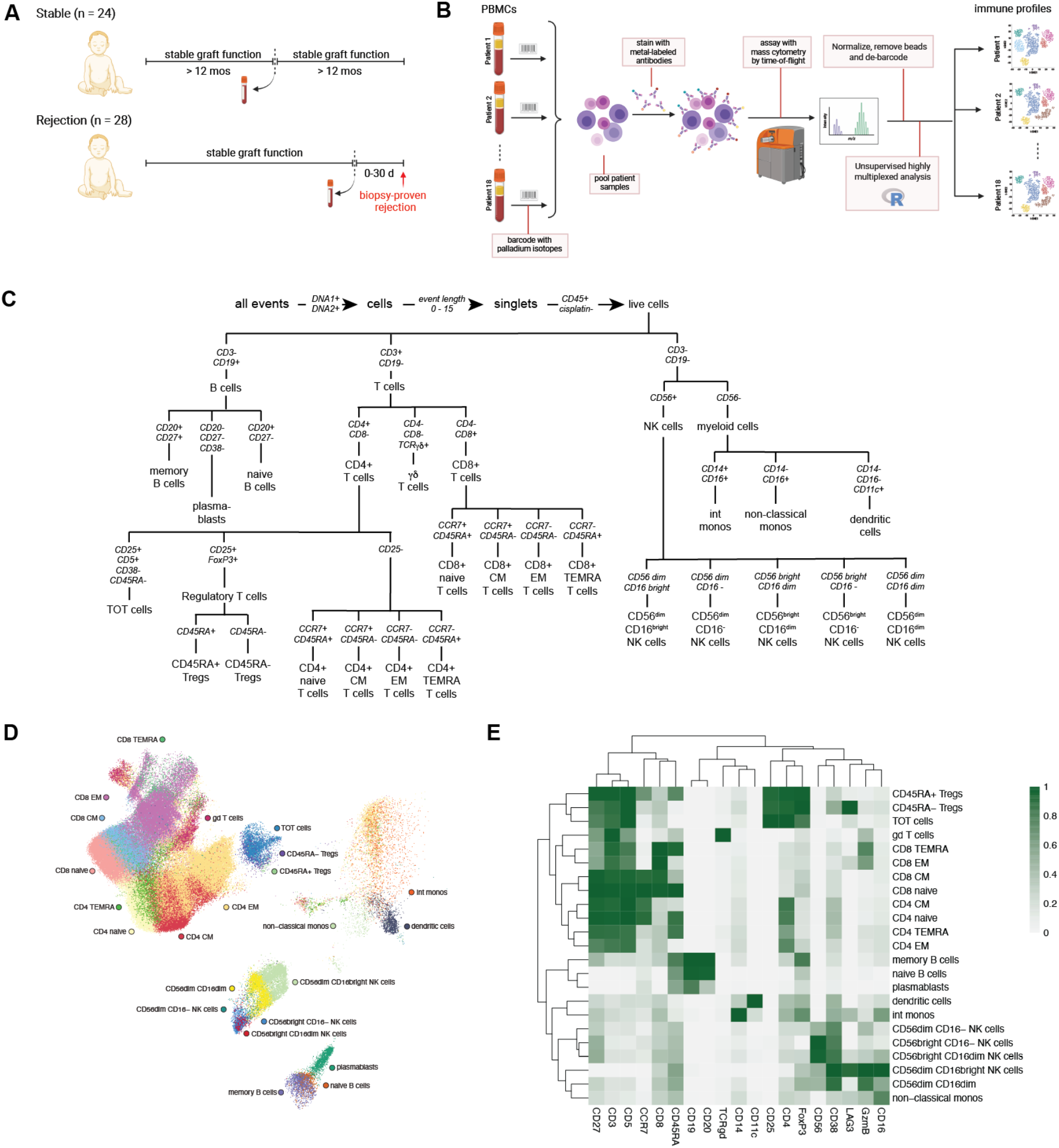
Experimental approach and sample processing workflow. A) Patient samples were obtained from a biobank constructed as a part of the Clinical Trials in Organ Transplantation-06 (CTOTC-06) study. Patients were classified as “stable” if they had no evidence of rejection for 12 months before and 12 months after sample collection; patients were classified as “rejection” if they developed biopsy-proven rejection 30 days or less after sample collection. B) PBMCs from donors were thawed, “barcoded” with palladium isotopes, stained with a combination of intracellular and extracellular markers, and analyzed on a mass cytometer. Normalized and debarcoded mass cytometer data was subject to both supervised and unsupervised analyses to generate immune profiles from patients, as described in **Methods**. C) For each patient sample, live singlet cells were identified based on DNA content, event length, and live/dead staining. Twenty-nine subpopulations of live singlets were identified using previously-published combinations of surface markers. D) The events in the terminally-differentiated branches of the tree outlined in (C) from all clinical samples were pooled and used to construct a single-cell UMAP (clustering markers CCR7, CD8, CD45RA, CD25, CD3 CD5, CD4, FoxP3, CD56, CD38, GzmB, CD16, CD19, CD20, CD14, TCRγδ, CD11c, CD25, LAG3) to show the phenotypic relationship between the terminally-differentiated populations. Cells are colored by their manually-gated population. E) Hierarchically clustered heatmaps visualize the relationship between populations and correlations between marker expression. The median expression of each marker used for gating was then assessed for each of the populations identified in (C).

### High-dimensional analysis identifies 29 different immune cell populations

Using an approach similar to that described in Neeland, et al.,^24^ manual gating was used to identify 29 subpopulations of PBMCs using the gating strategy described in **Figure 1C**. To determine if the relationship between the gated populations in high-dimensional space was consistent with known phenotypic similarities between cell types, the cells in the 23 terminally-differentiated branches of the tree outlined in **Figure 1C** from all clinical samples were pooled and used to construct a single-cell UMAP. UMAP facilitates the exploration of large, high-dimensional datasets by generating a two-dimensional (2D) coordinate system that enables simultaneous visualization of all cells in a single biaxial plot to capture the high-dimensional relationships of cells. UMAP spatial relationships were consistent within manual CellEngine gating. The major T cell, B cell, NK cell, and monocyte lineages separated in a biologically meaningful manner, and their cell subtype relationships were largely consistent with known biological patterns, such as the separation between lymphoid and myeloid lineages (**Figure 1D**). Hierarchical clustering of median marker signal intensity for each manually-gated population separated lymphoid and myeloid subsets and captured more granular cell type relationships between CD4 and CD8 T cells (**Figure 1E**).

### The post-transplant immune profile depends on the allograft type

The goal of this study was to understand those features of the immune response that lead to rejection that were common to multiple different allograft types. As such, our patient population included children who had received one of four different types of allografts, with either stable graft function or developing rejection. We used PCA, an unsupervised approach to exploratory data analysis, to better understand patterns present in the high-dimensional immune profiles across the entire cohort. We qualitatively observed that when grouping by allograft health alone, there was overlap between the groups, suggesting that the high-dimensional immune profiles were globally similar **(Figure 2A)**. When grouping by allograft type, the groups showed separation in principal component (PC) space (**Figures 2B, S1A, S1B)**. PC1 separates immune profiles of heart recipients from kidney and liver recipients, while PC3 separates kidney from liver recipients. No conclusion could be drawn regarding the immune profile of intestinal transplant recipients given the small number of samples included in the cohort. Quantification of the proportion of variance explained by allograft health and allograft type across the high-dimensional immune profiles using mixed models showed that the allograft type accounted for a greater proportion of the variance compared to allograft health (**Figure S1C**).

**Figure 2:**
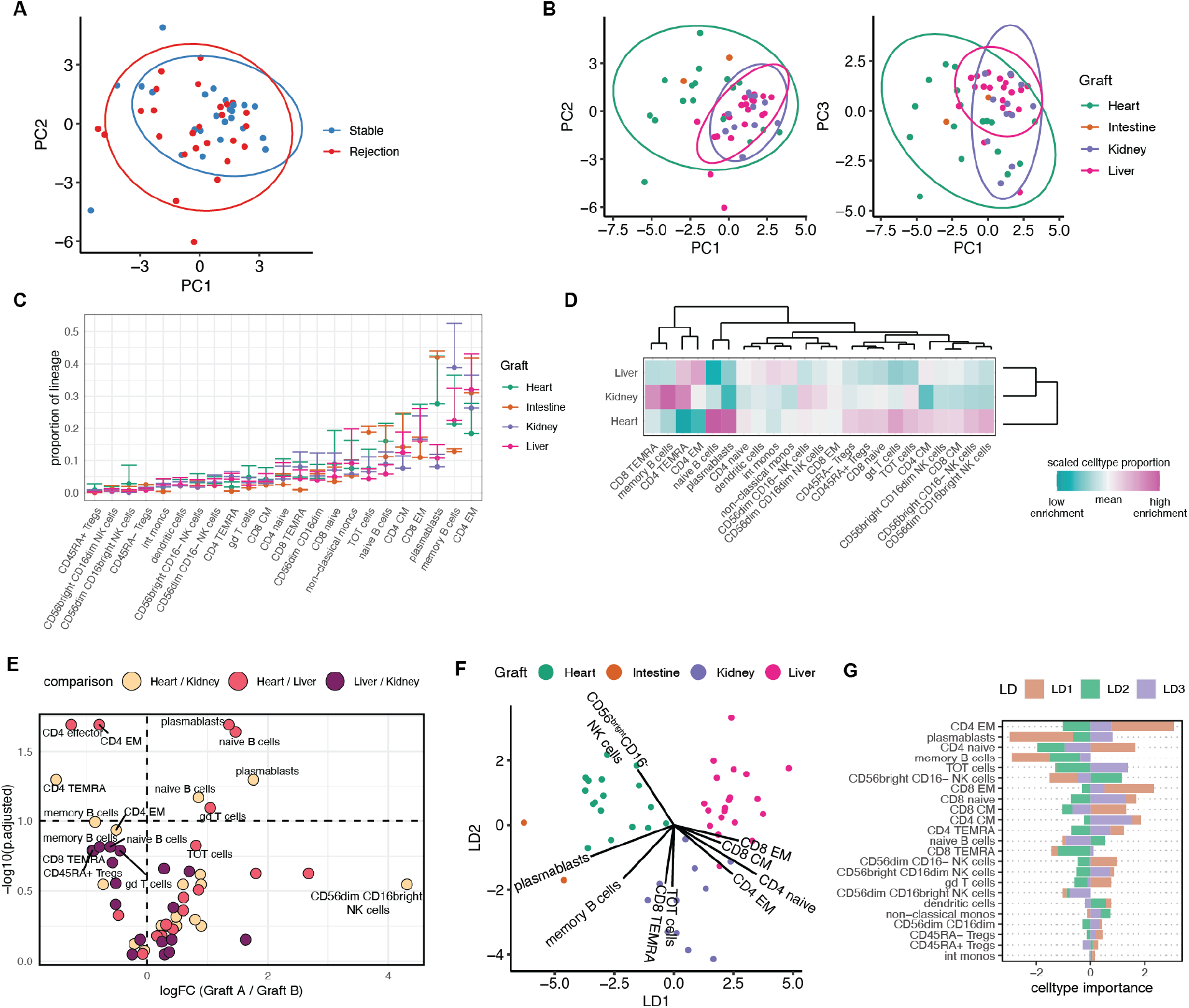
The post-transplant immune profile depends on the allograft type. A) Biaxial plot of PC1 and PC2, colored by graft health. B) Biaxial plot of PC1 vs. PC2 and PC1 vs. PC3, colored by graft. C) Proportions by lineage for each cell type across the different grafts. Error bar indicates standard deviation. D) Hierarchically-clustered heatmap of mean cell type proportions for each graft. E) Volcano plot summarizing results of differential proportions analysis between pairs of different grafts (excluding intestine). Cell type proportions compared using Wilcoxon signed-rank test, corrected for multiple hypotheses with an adjusted p-value cutoff at 0.1. F) Biaxial plot of LD1 and LD2 from LDA of immune cell type proportions by graft. Arrows indicate cell type importance for the top contributing immune cell types separating grafts along LD1 and LD2. G) Cell type importance results from LDA in (F) for LD1, LD2, and LD3.

Analysis of the mean and variance (standard deviation) between immune cell proportions for the 23 terminally-gated populations between the liver, heart, and kidney recipients (n = 21, 18, and 11, respectively) showed certain immune cell subpopulations had similar mean and variance in proportion across different allograft types (e.g., CD45RA+ Tregs and CD56^bright^ CD16^dim^ NK cells) while others varied significantly in their mean and variance, (e.g., plasmablasts, memory B cells, and CD4+ EM T cells, **Figure 2C**).To determine whether any of the immune cell subtypes were differentially abundant in an allograft-dependent manner, we analyzed immune cell proportions for 23 terminally-gated populations between liver, heart, kidney, and intestinal recipients (n = 21, 18, 11, and 2, respectively) **(Figure 2C)**. Since there were only two patients that had received intestinal transplants, we removed them from subsequent analyses. Hierarchical clustering of the mean cell type proportions between each allograft type showed that the immune compositions of liver and kidney recipients were more similar to each other than to heart recipients (**Figure 2D**). On average, peripheral blood from heart recipients had greater enrichment of naïve B cells, plasmablasts, and γδ T cells and decreased enrichment of CD4 TEMRA and CD4 EM populations compared to liver and kidney recipients. Peripheral blood from kidney recipients had higher enrichment of CD4 TEMRA, CD8 TEMRA, and memory B populations and decreased enrichment of plasmablasts and CD4 CM populations than the liver and heart recipients. The biologic basis for these allograft-specific signatures was not clear to us, so we looked at other confounding factors that may bias our findings. One possible explanation for these differences in immune composition is the effects of immunosuppression. We compared the immunosuppressive agents that patients were receiving at the time of sample collection (**Table 3**). Approximately 90% were receiving tacrolimus across all allograft types. Use of mycophenolate was variable across allograft types, with ∼75% of heart and kidney recipients but only ∼33% of liver recipients receiving mycophenolate at the time of sample collection, highlighting one possible difference in immunosuppression regimen that could contribute to differences in immune composition. However, we observed greater similarity between kidney and liver recipients compared to heart recipients, suggesting that use of mycophenolate is not sufficient to account for differences in immune profile. We also compared induction medications across allograft types (**Table 3**). Induction regimens differed across allograft types; however, given that, on average, samples were collected approximately two years after transplant, the residual effects of induction therapy are likely to be minimal. It has been well-established that the immune composition of a child changes with age, so another possible explanation for differences in immune composition that we observed could be that the age distribution of patients differed between allograft cohorts. Comparison of the demographics of the patients in each allograft cohort revealed that 1) each allograft cohort had patients from a wide variety of age groups, and 2) there were not significant differences in the age makeup of patients across cohorts (**Table 3**).

**Table 3:** Comparison of patient characteristics by allograft type

|  | Allograft Type |  |  |  |  |  |
| --- | --- | --- | --- | --- | --- | --- |
| Immunosuppression | Heart (18) | Kidney (11) | Liver (21) |  |  |  |
| Tacrolimus | 88.9% | 90.9% | 85.7% |  |  |  |
| MMF | 72.2% | 72.7% | 33.3% |  |  |  |
| Cyclosporine | 5.6% | 9.1% | 4.8% |  |  |  |
| Azathioprine | 5.6% | 0.0% | 0.0% |  |  |  |
| Cyclophosphamide | 0.0% | 0.0% | 0.0% |  |  |  |
| mTOR Inhibitor | 5.6% | 0.0% | 9.5% |  |  |  |
| Corticosteroid | 33.3% | 18.2% | 9.5% |  |  |  |
| Methylprednisolone | 0.0% | 9.1% | 0.0% |  |  |  |
| Other | 16.7% | 0.0% | 4.8% |  |  |  |
| Induction Medication |  |  |  |  |  |  |
| ATG (Thymoglobulin) | 33% | 55% | 10% |  |  |  |
| Basiliximab | 11% | 9% | 5% |  |  |  |
| Methylprednisolone | 22% | 36% | 52% |  |  |  |
| IVIg | 17% | 0% | 0% |  |  |  |
| None | 17% | 0% | 33% |  |  |  |
|  |  |  |  | p value |  |  |
|  |  |  |  | Heart vs.<br>Kidney | Heart vs.<br>Liver | Kidney<br>vs. Liver |
| Age at Collection (days) | 3962 ± 2345 | 3819 ± 2508 | 2928 ± 2033 | 0.985 | 0.335 | 0.543 |
| Time Post Transplant (days) | 693 ± 499 | 551 ± 315 | 614 ± 487 | 0.703 | 0.857 | 0.928 |

Given that we observed differences in the high-dimensional immune compartments of different allograft types, we performed statistical analyses to determine whether any immune cell subtypes were differentially abundant in an allograft-dependent manner. Specifically, we asked whether there were statistically-significant differences in the proportion of any immune populations between two graft types. Indeed, cells in the lymphocyte compartment were significantly differentially abundant in an allograft-dependent manner, even after multiple hypothesis correction using the false discovery rate (**Figure 2E**). Specifically, plasmablasts, naive B cells, CD4 TEMRA, CD4 EM, and γδ T cells were differentially abundant when comparing heart to liver recipients, while CD4 TEMRA, naïve B cells, and plasmablasts were differentially abundant when comparing heart to kidney recipients. Consistent with the hierarchical clustering, most cell type proportions were differentially abundant in the heart compared to the liver and kidney.

Correlation analysis of the immune cell compartments also showed that the correlative relationship between immune cell type proportions in each allograft were also different (**Figure S1D**). Specifically, we found that the magnitude and direction of the correlative structure between cell type proportions differed between allograft type. For example, among kidney recipients, the proportion of peripheral CD4 naïve cells was positively correlated with the proportion of CD4 CM, plasmablasts, and naive B cells; however, in heart or liver recipients, the abundance of these same populations was either more weakly positively correlated or negatively correlated.

Our initial differential abundance analysis comparing grafts in a pairwise manner found larger differences between the post-transplant peripheral immune compartment of heart recipients when compared to the liver and kidney recipients. Both the correlation analysis and PCA suggest that there may be differences in the immune compartment of the liver and kidney recipients as well. We performed supervised dimensionality reduction using LDA to further assess differences in the post-transplant peripheral immune compartment of these grafts by inputting the immune cell type proportions for all patients (**Figures 2F, S1E**). LDA is a classification algorithm that identifies linear combinations of features that optimally separate previously-determined class labels – such as allograft types – and has been used in both single-cell and clinical trial analysis.^22,25^ We exploit the inherent dimensionality reduction of LDA to visualize differences between samples. Indeed, the post-transplant peripheral immune compartment of each allograft type was strongly separated by LDA, corroborating that immune cell type proportions of patients with these grafts were indeed different. LDA reduces multiple cell types into linear discriminants (LDs), and each cell type is given a coefficient that has a magnitude and direction associated with the linear discriminant. The magnitude and direction of the coefficient indicates which cell types are the strongest drivers of differences between allograft types. LD2 adequately separates kidney from heart and liver recipients, while LD1 additionally separates heart from liver recipients. Consistent with our differential expression analysis, lymphocyte subpopulations were some of the strongest contributors to both LD1 and LD2, including naive B cells, CD4 TEMRA cells, CD4 EM cells, CD8 EM cells, and CD8 CM cells (**Figure 2G**). T cells of operational tolerance (TOT), a CD25+ CD5+ CD38-CD45RA-subset of CD4 T cells first reported in 2016 as upregulated in operationally-tolerant pediatric liver transplant recipients,^15^ were also strong contributors to LD2, suggesting that TOT cells may also have differential abundance in an allograft-dependent manner.

We additionally performed a follow-up random sampling analysis to ascertain whether this allograft-dependent result was due to random patient-level heterogeneity using sparseLDA, an optimized version of LDA for datasets with small samples sizes and a large number of features (eg, cell types). Indeed, sparseLDA of 1000 random samples analyzing three groups of equal size to the heart, kidney, and liver recipients failed to separate the 3 randomly sampled groups when compared to the ground-truth allograft analysis (**Figure S2**); thus, this allograft-dependent difference in the post-transplant immune compartment of these patients was not due to random patient-level heterogeneity. This comprehensive analysis using both unsupervised and supervised methods to study cell type proportions indicates that there are cell types in the post-transplant immune compartment of pediatric heart, kidney, and liver recipients that differ in an allograft-dependent manner.

### Rejection is associated with changes in proportion of distinct PBMC subpopulations

One goal of this study was to determine if there was an “immune signature” of allograft rejection, i.e., distinct changes in the proportions of peripheral blood mononuclear cell populations that were associated with the development of rejection. Unsupervised analysis by PCA showed no separation between stable and rejecting patients (**Figure 2A**). However, given that we saw allograft-level differences between many of these subpopulations, and that there are differences in immunogenicity across organs, we hypothesized that abundance of peripheral immune cells as markers of allograft stability or rejection may be masked by basal differences in cell type proportions between patients with allograft types. For example, if the proportion of a specific cell type is generally higher among heart recipients than other allograft types but also changes with allograft health, then a traditional statistical test comparing average cell type abundance in stable vs. rejecting patients will not capture this biology. Linear mixed models can be used to partition the variance of each feature (e.g, cell type) across experimental or clinical conditions, and summarize the contribution of each cell type in terms of the fraction of variation explained (FVE).^18,26^ To account for this, we applied linear mixed models to measure the variance of each cell type’s proportions associated with allograft type and allograft health in our cohort of transplant patients. We found that the variance of lymphocyte populations was associated with allograft type, allograft health, or both (**Figure 3A**). This was consistent with our prior differential abundance analysis, where the cell type proportion of several populations, including plasmablasts, CD4 EMs, CD4 TEMRA, and naïve B cells, were significantly different between allograft types (Figure 2E). Interestingly, the total variance in cell type proportions was more strongly associated with the allograft type than allograft health (**Figure S1C**). We also found that CD8 naïve, CD8 CM, and TOT cells were specific populations with the largest variance associated with allograft health relative to other cell types (**Figure 3A**). Hierarchical clustering of the mean cell type proportion across all allograft types and allograft health states also showed that the post-transplant peripheral immune compartments of different grafts were more similar within each allograft type regardless of allograft health status (**Figure 3B**). Interestingly, TOT cell abundance was associated with both allograft type and allograft health, and the average TOT cell proportion was higher in patients with stable graft function (compared to those experiencing rejection) within each allograft type. Given that CD8 naive, CD8 CM, and TOT cells were populations whose abundance was associated with allograft health, we performed differential abundance analysis between the stable and rejection cohorts in these three major populations. We implemented generalized linear models (GLMs) which have been commonly used in both clinical trial and single-cell analysis to compare the stable cohort to the rejection cohort, controlling for basal differences in cell type abundance between allograft types.^27–29^ The advantage of this approach is that it allows us to leverage all samples in the same statistical test, improving statistical power. The proportions of CD8 naive, CD8 CM, and TOT cells were all significantly associated with allograft health **(Figure 3C)**. Increased TOT cell proportion was positively associated with stable grafts, while increased CD8 naive and CD8 CM proportion was positively associated with rejecting grafts. We sought to further ascertain whether these cell populations were directly associated with allograft health using logistic regression models comparing the stable cohort to the rejection cohort. First, we performed a lasso-regularized logistic regression model comparing the stable to the rejection cohort across all gated populations (data not shown). The final cell types selected as important in reducing the prediction error of allograft health included CD8 naive and TOT cells, with coefficients of approximately -4 and +2, respectively. Classical monocytes were also selected as an important predictor, however, the coefficient was less than 0.1, suggesting that while monocytes were selected by the model, it added negligible predictive power to the model in comparison to other selected cell types. We also performed a leave-one-out-cross-validation (LOOCV) logistic regression model comparing the stable cohort to rejection cohort using only the CD8 naive, CD8 CM, and TOT cell type abundances that were significantly associated with allograft health (data not shown). We achieved 62.5% accuracy in correctly predicting stability or rejection. Predictions better than a coin flip (62.5% > 50%) suggests these populations as important for allograft health. These modeling results further support the importance of TOT cells as well as CD8 naive and CD8 CM abundance in determining allograft health in the post-transplant immune system of pediatric transplant recipients.

**Figure 3:**
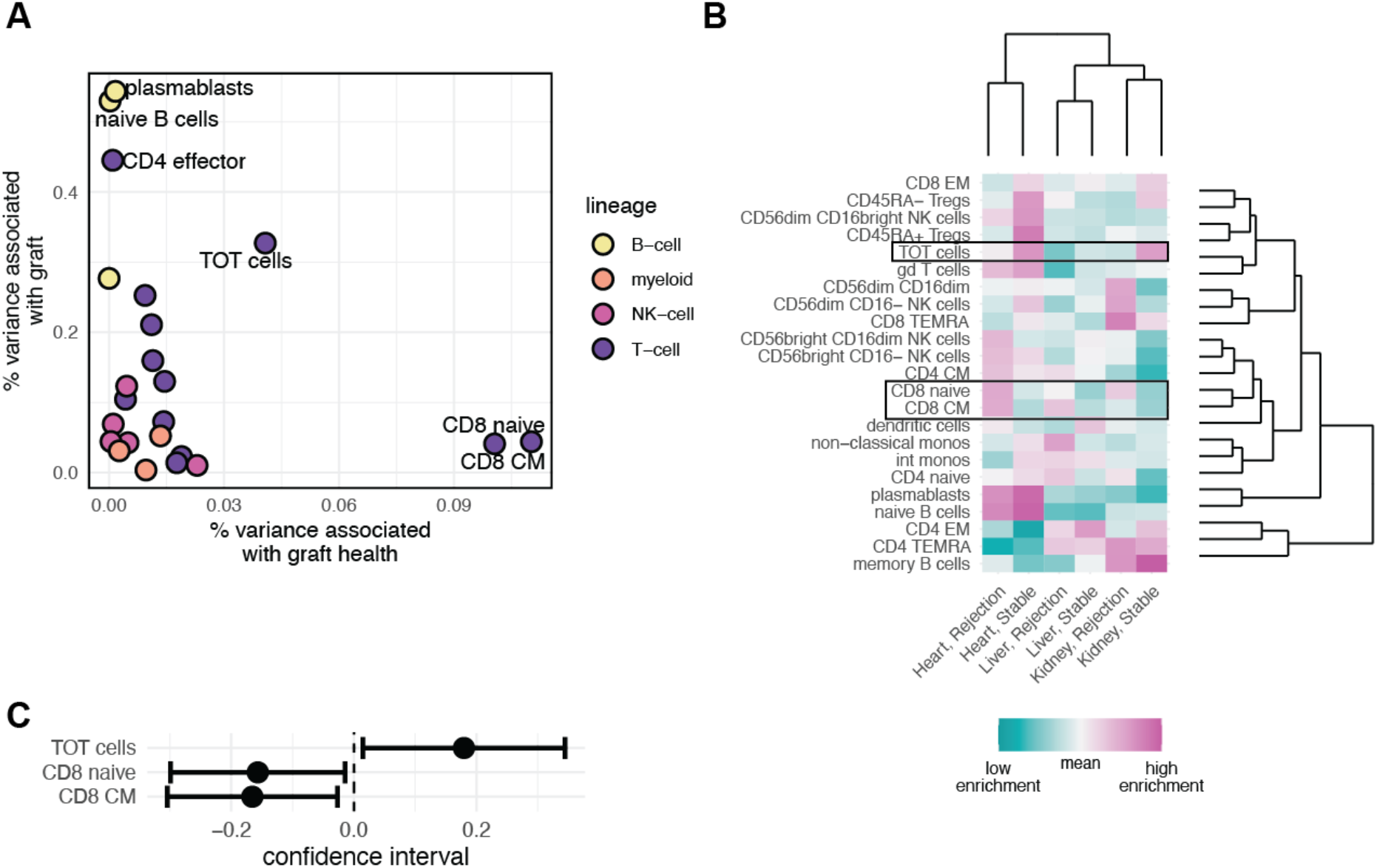
Rejection is associated with changes in proportion of distinct lymphocyte subpopulations. A) Biaxial plot showing results of linear mixed modeling analysis to calculate the fraction of variance explained by graft and graft health. Color indicates cell type lineage. B) Hierarchically clustered heatmap of mean cell type proportions across each clinical subset of graft and graft health. Boxes highlight key populations from (A). C) Confidence intervals for statistically comparing cell type populations that vary with graft health using generalized linear models: p <= 0.05.

### TOTs have a phenotype between that of effector T cell and regulatory T cell subpopulations

We had previously observed that TOT cells are more abundant in operationally-tolerant liver transplant patients, i.e., liver transplant patients who maintain stable allograft function even while off immunosuppression.^15^ This raises the possibility that TOTs have some inherent immunosuppressive function similar to regulatory T cells (Tregs). Because of the small number of cells available in these patient samples, isolation of TOTs for *in vitro* functional characterization was not feasible. As an alternate means to gain insight into the identity of TOTs, we compared the phenotype of TOTs to the six other CD4+ subpopulations queried in this study (**Figure 1C**) defined by the gating strategy shown in **Figure 4A**.^30^ The events in the seven CD4+ subpopulations from all clinical samples were pooled and used to construct a single-cell UMAP with the subset of markers in the panel known to be expressed on CD4+ T cells faceted by each of the seven CD4+ subpopulations (**Figure 4B**). TOTs were spatially proximate to the CD45RA-Tregs, indicating phenotypic similarity to this subpopulation. We then examined expression of each of the CD4+ T cell markers used to construct the UMAP across all the CD4+ populations. TOT cells had high expression of CD25 and CD45RO, low expression of FoxP3, and did not appear to express CD45RA, PD-1 or CD38 (**Figure 4C**).

**Figure 4:**
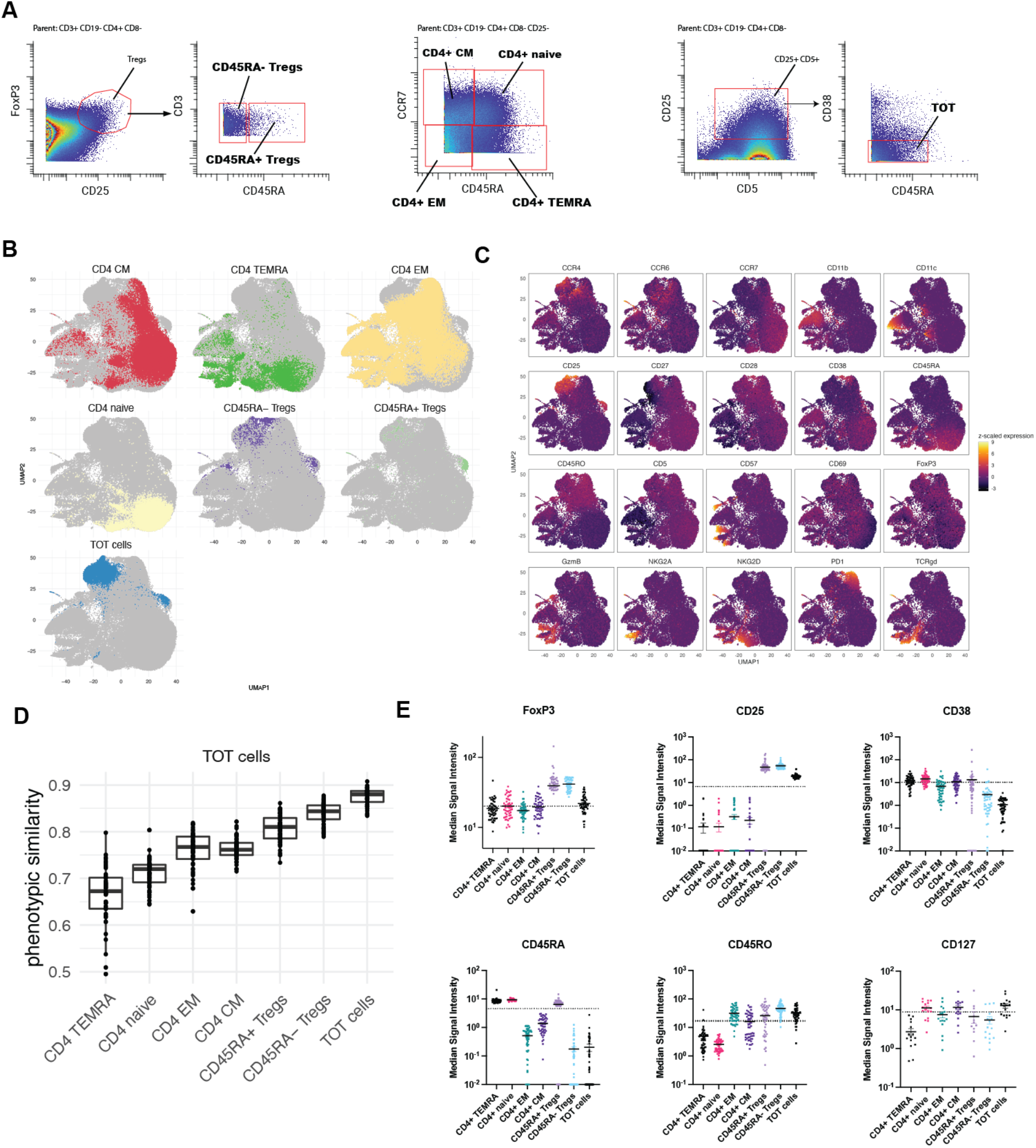
TOT cells have phenotypic similarities to regulatory T cells. A) Gating strategy used to define key subpopulations of CD4+ T cells. B) The events in the seven CD4+ T cell subpopulations outlined in (A) from all clinical samples were pooled and used to construct a single-cell faceted UMAP using markers known to be expressed on CD4+ T cells (CCR4, CCR6. CCR7, CD11b, CD11c, CD25, CD27, CD28, CD38, CD45RA, CD45RO, CD5, CD57, CD69, FoxP3, GzmB, NKG2A, NKG2D, PD1, and TCRγδ) to show the phenotypic relationship between these CD4+ subpopulations. C) A faceted UMAP using the same events in B) shows the expression levels of various T cell markers. D) Cosine similarity analysis of CD4+ subpopulations identified in A) E) The median signal intensity of selected T cell markers for each of the CD4+ subpopulations identified in A).

To quantitatively measure the phenotypic similarity between TOTs and the other CD4+ populations, we performed cosine similarity analysis.^31^ High-dimensional phenotypic (dis)similarity was estimated as a cosine distance to an average cell in each CD4+ population for each sample. Populations that are similar to each other will have a cosine similarity metric closer to 1, and those that are dissimilar will have a cosine similarity metric closer to 0. The TOT populations across all the clinical samples were most similar to themselves (average cosine ∼0.87). The populations that they were next most like (average cosine ∼0.83) were the CD45RA-Tregs, and the populations that they were least like (average cosine ∼0.65) were the CD4+ TEMRAs (**Figure 4D**).

We next compared the signal intensity of selected markers relevant to regulatory function or T cell activation between TOT populations and other CD4+ populations across all clinical samples. On average, TOTs express higher levels of two key regulatory T cell markers - FoxP3 and CD25 - than other CD4 naïve, CM, EM, and TEMRA cells but not as high as CD45RA+ and CD45RA-Treg cells (**Figure 4E**). Other T cell markers – CD45RA, CD45RO and CD38 – were expressed at similar levels between TOTs and CD45RA-Tregs. Lastly, like CD4+ naïve and CM subsets, TOTs have low level expression of CD127; this is distinct from CD45RA+ and CD45RA-Tregs, which are CD127-. Together, these data suggest that TOTs are most phenotypically similar to the CD45RA-subset of Tregs but have distinct features that make it unlikely that they merely represent a subset of this population.

## Discussion

In this study, we used mass cytometry (CyTOF) to construct multi-lineage immune profiles from a large group of pediatric heart, liver, kidney, and small bowel transplant recipients. To our knowledge, this is the first in-depth analysis of peripheral blood immune composition in pediatric transplant recipients. Much of the previous literature on peripheral blood immune composition of pediatric transplant recipients has been limited to single-lineage or has had limited depth within each subset.^32–34^ This is largely due to technical limitations, and the use of a 37-marker mass cytometry panel allowed a much more granular characterization at the immune composition than has been previously described.

One of the significant findings that emerged from analysis of these data is that the immune composition of the peripheral blood following solid organ transplant is correlated with the type of allograft that the child received, with the largest separation occurring between heart recipients and liver or kidney recipients. The reason for this phenomenon is not completely understood. One possible explanation is that these differences existed prior to transplant. There is some evidence in the literature to support the idea that heart failure, one of the leading indications for heart transplant, is associated with alterations in immune function. For example, patients with heart failure had a higher abundance of natural killer subsets that expressed granzyme B or fractalkine receptor compared to healthy controls; these patients also had a higher proportion of conventional dendritic cell populations and a lower proportion of classical monocytic populations. Interestingly, no significant changes in the abundance of major T or B cell populations were observed.^35^ If changes in immune function led to heart failure and these changes persisted after transplant, one might expect the heart failure to recur rapidly, which is not supported by clinical data. Many of the recipients profiled in this study are months or years from transplant, and any effect of organ failure on immune composition should have resolved. Another possible explanation is that thymectomy performed at the time of heart transplant may result in alterations in immune function. It has been reported that children who undergo thymectomy at the time of transplant have lower percentages of total Tregs and naïve T regs but higher percentages of memory CD4+ cells; these patients are also more susceptible to the development of atopic conditions.^36^ However, the majority of thymic involution occurs by age two, so thymectomy is not necessary for older children. The patients profiled in our study had a mean age around seven and included toddlers, school-age children, and adolescents. The majority of these children, therefore, would not have required thymectomy at time of transplant. The distinct features of the immune composition that we observed in heart transplant recipients spanned a much broader age range than could be explained by the effects of thymectomy alone. A third possible explanation is that the observed allograft-specific differences result from differences in immunosuppressive treatment during induction or at the time of sample collection. As described above, the similarities in immunosuppressant use between allograft types were not correlated with the similarities in immune composition. Individual patient data on immunosuppressant target levels was not available to us, so the effects of different levels of immunosuppression (i.e., high tacrolimus levels vs. low tacrolimus levels) on immune composition cannot be excluded. Given the relative standardization of immunosuppressive regimens for a given allograft type across transplant centers, it is likely not feasible to completely separate the effects of immunosuppression from allograft type on immune composition. We also did not detect significant differences in age of patient or time from transplant across allograft cohorts. Determining whether differences in immune composition exist prior to transplant would give additional insight into this phenomenon and is the subject of an ongoing investigation.

If the immune response to an allograft is dependent on the type of the graft received, these differences may have significant implications for the development of rejection. There is some evidence in the literature consistent with idea that rejection may have organ-specific properties. For example, rates of rejection vary dramatically depending on allograft type, with the lower rates of rejection occurring in kidney (14%) or liver (27%) transplant recipients and higher rates occurring in heart or intestinal transplant recipients (40%). Additionally, operational tolerance – a state wherein allograft health can be maintained without administration of exogenous immunosuppression – has been observed in kidney and liver transplant recipients but only anecdotally in heart or intestinal transplant recipients.^37–39^ We also observed that the peripheral blood immune composition of kidney and liver recipients were more like each other than either was to heart recipients. The presence of organ-specific differences in post-transplant immune composition highlights the importance of first performing organ-specific analyses when looking at populations important for graft health.

When these organ-specific differences were considered, we observed that there were distinct subpopulations of T cells whose abundance in the peripheral blood was associated with graft health. First, we observed that increased frequency of CD8 naïve and CD8 CM populations were associated with the development of allograft rejection. A role for CD8 T cells in the pathogenesis of allograft rejection is well-established, but the roles of specific CD8 subpopulations are less well-defined, particularly in pediatric transplant recipients. On the one hand, there is evidence that highlights a role for CCR7+CD8+ populations – which include naïve CD8 and CD8 CM populations – in the prevention of rejection. In an in vitro study, CCR7+ CD8+ cells from healthy humans suppressed the proliferation of and production of IL-2 and IL-17 by CD4+ T cells stimulated by addition of anti-CD3 and anti-CD28 antibodies.^40^ However, the same study also observed a higher abundance of CCR7+ CD8+ populations in the peripheral blood of adult kidney transplant recipients with T cell-mediated rejection compared to those with normal biopsies.^40^ Another study that examined the composition of the peripheral immune compartment in adult kidney transplant recipients reported higher percentages of naïve CD8 T cells in patients experiencing biopsy-proven late rejection; a similar trend was seen in the abundance of CD8 CM T cells.^41^ The relationship between abundance of naïve CD8 T cells, CD8 CM T cells and allograft health demonstrated here further underscore the importance of these cell types in the pathogenesis of rejection.

We also observed that increased frequency of TOT cells was associated with stable allograft function. There is limited data on function and phenotype of TOT cells in the literature. TOTs were first reported as a subpopulation of CD4+ T cells in 2016 using machine learning approaches to identify novel combinations of surface markers expressed on PBMCs in pediatric liver transplant patients.^15^ Importantly, combination of cell surface markers expressed by TOTs had not previously been described in the literature and was only identified by using many cellular markers in combination with unsupervised analyses. This highlights the importance of performing high-dimensional analyses on patient samples and interrogating the data with unsupervised algorithms to truly appreciate the complexity of the immune response to an allograft.

TOTs make up a significant portion of the CD4+ compartment (5-10%); a higher frequency of TOTs is associated with operational tolerance, the ability of an individual to maintain stable allograft function without exogenous immunosuppression, in pediatric liver transplant recipients.^15^ The data from this study suggests a much larger role for TOT cells in maintenance of allograft stability, as we observed that increased frequency of TOT cells was associated with graft stability in heart, liver and kidney recipients. Another important distinction between this study and what we observed previously is the effect of immunosuppression on TOT abundance. It had been previously reported that TOTs were upregulated in patients who were not receiving immunosuppression;^15^ here, we observe a similar degree of upregulation of TOTs even in patients receiving maintenance immunosuppression. This result from the current study suggests that 1) TOT cells are not negatively regulated by classical forms of immunosuppression and 2) augmentation of TOT abundance or function may represent an alternative route to maintenance of allograft stability outside of that mediated by classical immunosuppressive agents. Furthermore, since the development of rejection was associated with changes in frequency of TOTs, they may represent novel biomarkers for the non-invasive detection of rejection.

It is important to note that many of the differences in immune cell abundance between stable and rejecting patients were not apparent when the effects of allograft type were not taken into account. This suggests that previous studies of differences in immune composition that did not take the organ-specific differences into consideration may have failed to detect differential abundance in populations between stable and rejecting patients. In our dataset, we also noted that there were other cell types with 0.5 to 1 log fold-change differences between stable and rejecting patients, but these differences were not significantly different. Due to wide patient-level heterogeneity and small sample size, it is possible that other immune cells may be differentially abundant that were not captured in this pediatric trial.

We also demonstrated that TOT cells were phenotypically most similar to CD45RA-Tregs but have enough differences in expression of key markers that it is unlikely that they are merely a subpopulation of these cells. Furthermore, TOTs are several-fold more abundant in the peripheral circulation than the CD45RA-Tregs. This suggests that TOT cells may represent a “ready reserve” of CD4+ T cells that can be induced to suppress immune activation when needed. An equilibrium between effector and regulatory T cell populations has been described previously in the literature, with multiple cytokine-signaling pathways implicated in driving the balance in one direction or the other. For example, TGF-β, an anti-inflammatory cytokine, induces FoxP3 and the development of Treg cells in the periphery; however, the combination of the pro-inflammatory cytokine IL-6 and TGF-β induces generation of effector Th17 populations and TGF beta is required for this process.^42^ The transcription factors RORγt/RORa (found in Th17 populations) and FoxP3 (found in Treg populations) can bind and inhibit each other’s function.^43,44^ IL-2 promotes growth of Tregs and inhibits Th17 differentiation, while IL-21 promotes Th17 differentiation and inhibits Treg expansion.^45^ If TOTs indeed represent a “ready reserve” of CD4+ T cells with regulatory capabilities, the above data suggest that we may be able to “nudge” them towards a regulatory phenotype with exogenous cytokine stimulation as a novel therapy for prevention or treatment of rejection.

One potential pitfall in analyzing these data is that clinical evidence of rejection may lag behind the immune events that accompany rejection. In the absence of a biopsy that clearly demonstrates that immune infiltration of the allograft has not occurred, one cannot be sure that there is not a low-level immune activation occurring even when the patient has no clinical evidence of rejection. To minimize the potential of misclassifying a patient with low-level rejection as stable, we stipulated that patients should not have any evidence of rejection for the 12 months after sample collection in order for it to be used for analysis in the study. We reasoned that meaningful immune activation leading to rejection would manifest in the 12-month period after sample collection and therefore those patients who maintained stable graft function for 12 months after sample collection were unlikely to have low-level rejection at the time of collection.

In summary, we have demonstrated that the immune response to an allograft was strongly associated with the type of allograft received and increased abundance of TOT was strongly associated with graft stability across multiple different types of allografts. Because a limited set of cell-surface markers can be used to define TOT cells, their abundance can be measured in individuals using clinical flow cytometry-based assays, and this may represent a novel, minimally-invasive approach to monitor for the development of allograft rejection. Measurement of TOT abundance in transplant recipients over time will need to be performed to determine whether there is an absolute threshold for TOT abundance that predicts rejection or whether changes in TOT abundance from pre-transplant or early post-transplant levels can be used to predict the development of rejection. Furthermore, because TOTs have features of regulatory cells, they may represent a population that can be targeted *in vivo* to selectively dampen an immune response in a patient experiencing rejection.

## Data Availability

All data produced in the present study are available upon reasonable request to the authors.

## Acknowledgements/Funding

This study was funded by awards from the NIH (UO1 AI104342 [C.O.E.], U01 AI1359590 [S.M.K.]), the Stanford Maternal and Child Health Research Institute [M.R.], the Stanford Transplant and Tissue Engineering Center of Excellence [M.R.] and the Stanford University Jackson Vaughan Critical Care Research Fund [M.R.]. M.A. is supported by the Stanford Immunology Training Grant T32 AI007290_37. The authors would like to extend their special thanks to all of the investigators in the CTOTC-06 consortium for their work in enrolling subjects in the parent study and providing samples for this investigation.

## Data Availability Statement

The data for this study will be available from the corresponding author upon reasonable request.

## Code Availability Statement

The code for this study will be available from the corresponding author upon reasonable request.

