## Supplemental Figures for "High-dimensional profiling of pediatric immune responses to solid organ transplantation"

### Figure S1

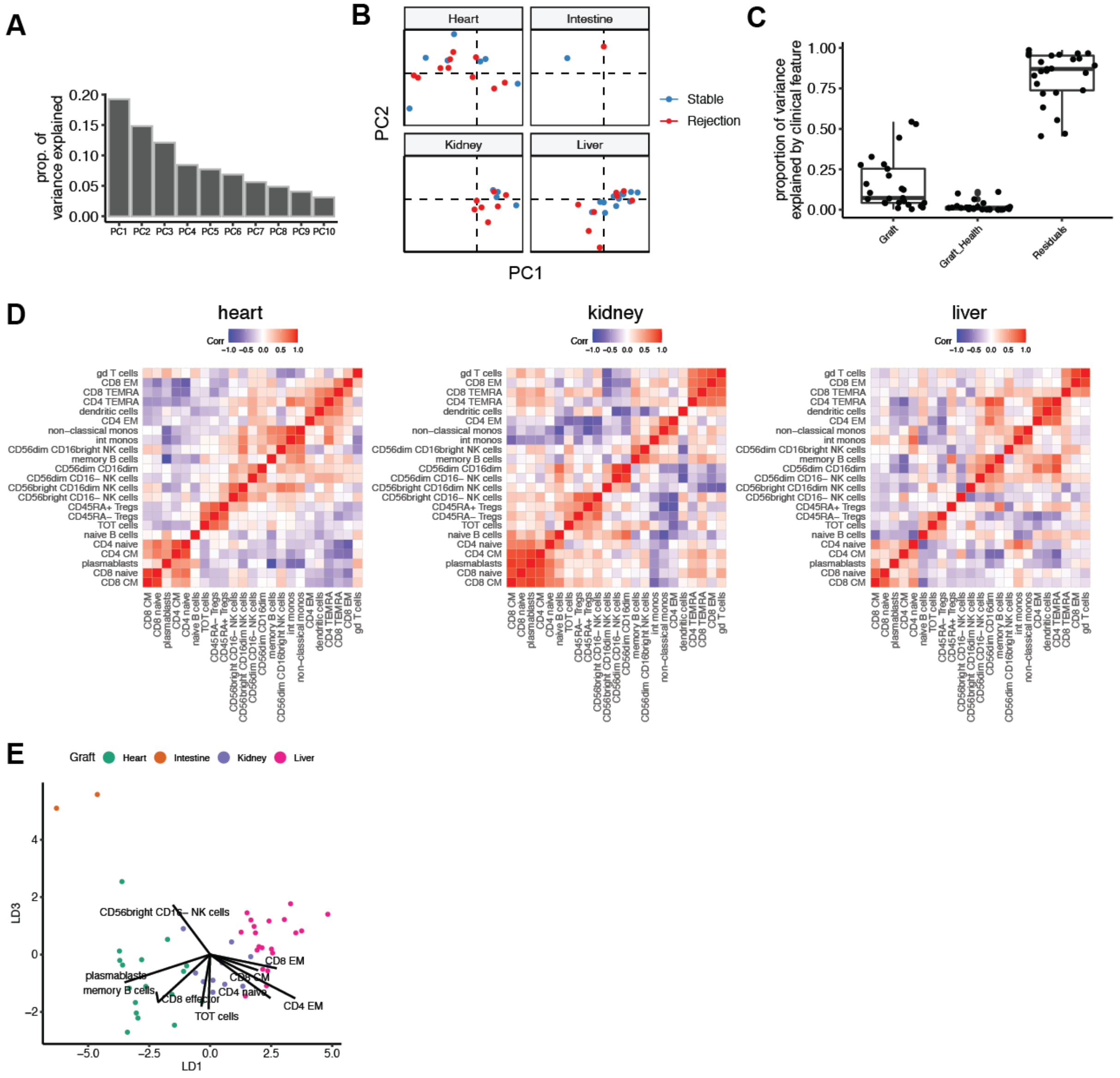

**Supplementary Figure 1: The relationship between the post-transplant immune profile of different allograft types.** A) Barplots of the proportion of variance explained by each PC from PCA. B) Biaxial plot of PC1 and PC2, colored by graft health and stratified by graft. C) Boxplot results of linear mixed modeling analysis showing the fraction of variance explained by graft and graft health for each cell type. D) Correlation plots of heart, kidney, and liver. Hierarchical clustering was performed on heart, and the same ordering was applied to kidney and liver to assist with comparing correlation maps. E) Biaxial plot of LD1 and LD3 from LDA of immune cell type proportions by graft. Arrows indicate cell type importance for the top contributing immune cell types separating grafts along LD1 and LD3. Cell type proportions are mean-centered and scaled prior to LDA.

### Figure S2

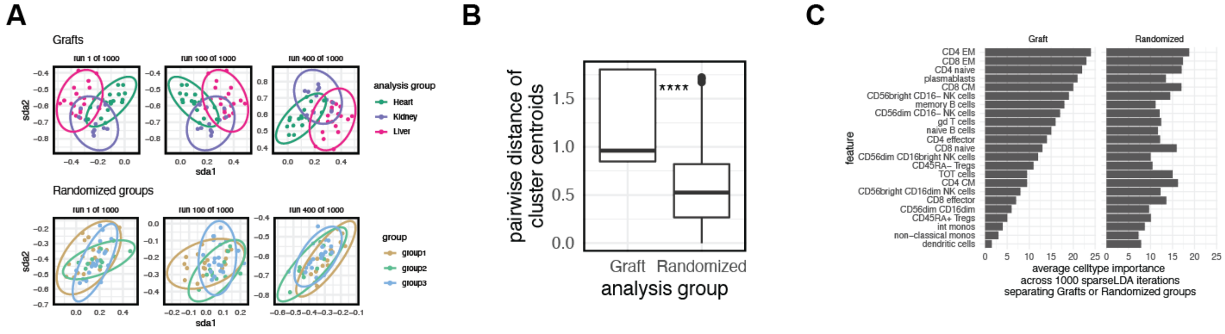


**Supplementary Figure 2: The relationship between the post-transplant immune profile of different allograft types is abolished with randomization.** A) Three random examples of sparse discriminant analysis results from the 1000 random permutations comparing cell type proportions across the grafts or randomly scrambled groups. Cell type proportions are mean-centered and scaled prior to sda. B) Pairwise distance of centroids in sda1 and sda2 for each of the three grafts compared to the 1000 randomly scrambled permutations. Mann-Whitney test **** = $p < 0.0001$. C) Average cell type importance determined by sda coefficients from 1000 sda iterations comparing graft or the randomly-scrambled group.

**Figure S3**

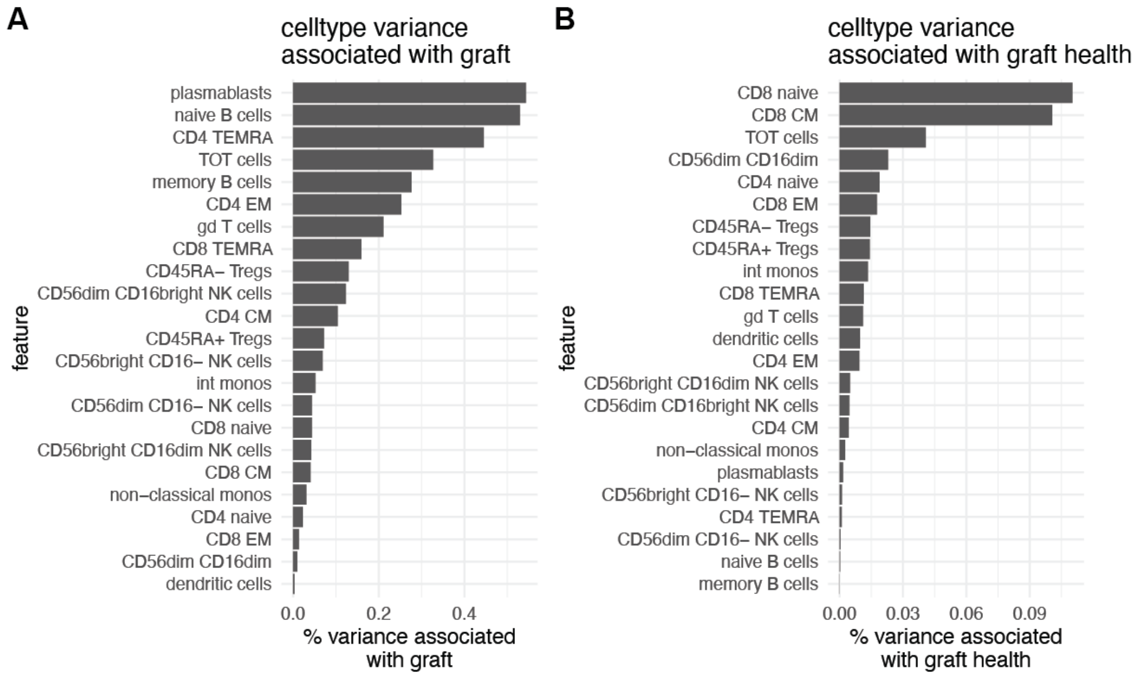


**Supplementary Figure 3: Summaries of cell type variance based on graft type and graft health.** Percent of variance associated with graft type (A) and graft health (B), for each cell type. Cell types ordered from largest to smallest percent of variance explained.
